# Distinct gut microbiota profiles may characterize amyloid beta pathology and mild cognitive impairment

**DOI:** 10.1101/2024.05.01.24306673

**Authors:** Konstantinos Rouskas, Eirini Mamalaki, Eva Ntanasi, Marianna Pantoura, Maria Anezaki, Christina Emmanouil, Nil Novau-Ferré, Mònica Bulló, Antigone S Dimas, Christopher Papandreou, Mary Yannakoulia, Anagnostis Argiriou, Nikolaos Scarmeas

## Abstract

Gut microbiome composition has been associated with early preclinical Alzheimer’s disease (AD), as reflected by cerebrospinal fluid (CSF) amyloid beta pathology, and with mild cognitive impairment (MCI). However, the presence of distinct microbiota across different disease stages has not been fully characterized. We profiled gut microbiota in 50 nondemented individuals by 16S ribosomal RNA sequencing and taxonomic profiles were compared between amyloid-based (amyloid-normal vs. amyloid-pathology) and clinically-based (cognitively normal vs. MCI) diagnosis groups using linear models (adjusted for sex, age and diet). Elastic net regression model was used to assess the discriminative performance of microbiota for amyloid-pathology and MCI. Microbial diversity measures did not differ across groups. We identified specific genera associated with amyloid-pathology and MCI such as *Oxalobacter, Marvinbryantia* and *Escherichia-Shigella*, mostly linked to inflammation. Distinct genera were found to be unique to amyloid-pathology and MCI. Microbiota was shown to have a fairly good discriminative performance. Overall, we suggest the presence of distinct microbiota in early preclinical stage of AD and MCI, which needs to be further explored.

## 1. Introduction

Alzheimer’s disease (AD)^1^ is the most common cause of dementia worldwide and a complex neurodegenerative disorder with significant burden for the society and the economy (Knopman et al. 2021). The diagnosis of AD is based on clinical criteria complemented with the biomarker-driven amyloid/tau/neurodegeneration (AT(N)) classification schema (Jack et al. 2018). This ATN framework characterizes the AD continuum by its biological presentation independently of clinical assessment of cognitive status. AD continuum begins from preclinical stages characterized by normal cognition and abnormal brain biomarkers, including amyloid-beta 42 (Aβ_42_), to mild cognitive impairment (MCI) and then to clinically apparent dementia (Tahami Monfared et al. 2023). Given that cerebrospinal fluid (CSF) Aβ_42_ pathology has been consistently associated with clinical progression to cognitive decline (Dumurgier et al. 2017; Prosser et al. 2023; Tijms et al. 2017), understanding of the distinct molecular alterations of early preclinical stages, compared to MCI, is imperative to earlier diagnosis and may serve for potential treatment strategies.

Accumulating evidence suggest that gut microbial dysbiosis is involved in AD pathogenesis probably via the gut-brain axis and by regulating peripheral and central inflammation through microbial metabolites (Chandra, Sisodia, and Vassar 2023). Recently, transplants of gut microbiota from patients with AD were shown to induce memory impairments in young animals (Grabrucker et al. 2023). Given that brain aggregation of amyloid Aβ_42_, the core pathology of AD (Knopman et al. 2021), seems to precede the development of significant cognitive decline and dementia (Insel et al. 2019), it is likely that the contribution of the gut microbiome to disease also occurs before symptom onset. To date, little information is available on the role of the gut microbiome in these early preclinical stages of AD (Verhaar et al. 2021; Jung et al. 2022; Ferreiro et al. 2023). Given that amyloid deposition, although not determinant, may lead to the development of the clinical signs of the disease, including MCI and AD dementia (Jack et al. 2018), it will be key to define microbes that distinguish individuals with MCI from cognitively healthy controls. Several lines of evidence suggest a role of gut dysbiosis in the development of MCI (Chen et al. 2023; Fan et al. 2023; Yildirim et al. 2022) however, according to our knowledge, there are no microbiome studies exploring both amyloid pathology and clinical status in a single population sample. Profiling microbial communities in both early preclinical and clinical stages of the disease can reveal distinct microbiota that may be used as early detection markers and potential treatment targets for AD.

Prediction models have a prominent role in healthcare research and clinical practice, as they can help physicians identify patients at risk of developing a disease and then recommend treatment plans (Eloranta and Boman 2022). Given that several lines of evidence suggest a role for gut microbes in the evolution of AD pathogenesis, it will be key not only to define differential gut microbiota across stages but also to investigate their discriminative capacity for amyloid pathology or MCI status. To date, little is known about the contribution of gut microbiome features in prediction models for amyloid pathology, highlighting microbes that produce short chain fatty acids (SCFAs) (Verhaar et al. 2021) or induce inflammation (Ferreiro et al. 2023), as the top taxonomic predictors. Given that CSF biomarkers are of limited clinical application due to discomfort during the lumbar puncture and/or high cost, the identification of low-cost and less invasive biomarkers, such as fecal microbiota, in further preclinical AD cohorts, would increase their use in clinical practice. Furthermore, identification of specific bacteria reflecting pathophysiological processes would help to identify biomarkers for AD pathology and MCI and contribute to further designing potential strategies to slow the rates of cognitive decline.

To this end, we performed 16s ribosomal RNA (rRNA) amplicon sequencing to profile and compare the gut microbiota composition between individuals with and without amyloid pathology and between cognitively normal individuals and those clinically diagnosed with MCI. We then investigated the presence of distinct microbiota alterations in amyloid-based and clinically-based comparisons. We further determined whether microbiota could discriminate for amyloid pathology and MCI and we investigated whether specific microbiota correlated with established AD biomarkers.

## 2. Methods

### 2.1 Study population

This research has been conducted within the ongoing ALBION (Aiginition Longitudinal Biomarker Investigation of Neurodegeneration) study initiated in 2018. The ALBION study sample consisted of individuals aged >40 years old, referred to the cognitive disorders’ outpatient clinic of Aiginition Hospital (Athens, Greece). Patients with a dementia diagnosis were excluded from ALBION, as well as patients with medical conditions associated with a high risk of cognitive impairment or dementia (including Parkinson’s disease, multiple sclerosis, hydrocephalus, epilepsy, Huntington’s disease, Down syndrome, active alcohol or drug abuse or major psychiatric conditions such as major depressive disorder, schizophrenia, and bipolar disorder). The baseline evaluation was completed by 198 individuals from 2019 to 2023. In the present study, we included only ALBION participants in their baseline evaluation with available fecal samples, CSF biomarker results and screened for MCI diagnosis (n=53) (**Fig. S1**). Among these, three individuals were excluded due to antibiotics use within three months prior fecal sampling. A detailed description of ALBION’s study protocol can be found elsewhere (Kalligerou et al. 2019; Scarmeas et al. 2022). Written informed consent was obtained from all participants, and study procedures were approved by the Institutional Review Board and Ethics Committee of the Aiginition University Hospital, National and Kapodistrian University of Athens, Greece (Protocol code: 255, ΑΔΑ: ΨΘ6Κ46Ψ8Ν2-8ΗΩ, date of approval: 10 May 2022).

### 2.2 Cerebrospinal Fluid (CSF) collection and analysis

All participants underwent comprehensive neuropsychological assessment by a certified neurologist. CSF was collected through lumbar puncture and stored following international guidelines (Teunissen et al. 2014). CSF was processed for biomarkers such as amyloid-β 1– 42 (Aβ_42_), phosphorylated tau at threonine 181 (pTau), and total tau (tTau). More details about CSF collection and analysis in ALBION can be found elsewhere (Kalligerou et al. 2019; Sampatakakis et al. 2023; Scarmeas et al. 2022). CSF samples were analyzed using automated Elecsys assays (Roche Diagnostics). The reference ranges for a positive result (pathology) were as follows: Aβ_42_ ≤ 1000 pg/ml, pTau >27 pg/ml and tTau >300 pg/ml. Given the low number of studied individuals having tau pathology (7 out of 50 individuals) and that amyloid aggregation is the first event related to neuropathology of AD (Zhang et al. 2018), possibly starting earlier than the accumulation of Tau (Wegmann, Biernat, and Mandelkow 2021), we restricted our analysis only on the amyloid status. Twenty-one out of 50 participants were classified as having amyloid-pathology, while the remaining 29 individuals were classified as amyloid-normal. We also categorized individuals based on established clinical diagnosis criteria (Petersen et al. 2001) to cognitively normal (CN) (n=40) and those having MCI (n=10).

### 2.3 APOE ε4 status and lifestyle data

*APOE* ε4 genotyping procedures have been described previously (Sampatakakis et al. 2023). Briefly, *APOE* ε4 genotyping was performed using a commercial kit (LightMix TIB MOLBIOL) in Roche Light Cycler 2 apparatus and hybridization probe method. Participants were classified as *APOE* ε4 carriers (at least one copy of the *APOE* ε4 gene) and *APOE* ε4 non-carriers (no copies of the *APOE* ε4 gene). Dietary intake was assessed by four 24-h recalls as described in a previous work (Brikou et al. 2023) and adherence to Mediterranean diet was assessed using an eleven-item composite score, the Mediterranean Diet Score (MDS) (Panagiotakos, Pitsavos, and Stefanadis 2006). Smoking status (former or present) was also assessed for participants.

### 2.4 Fecal sample collection, DNA extraction, library preparation and sequencing

Fecal samples were collected for all participants from November 2020 to January 2022. Each study participant was given a fecal collection container to collect a fecal sample at home. The participants were asked to store the sample in a freezer and to transport the samples to the hospital in a cooling bag the day following home collection. Fecal samples were then immediately stored at −80°C until DNA extraction. Microbial DNA samples were extracted using a QIAamp DNA Stool Mini Kit (Qiagen, Hilden, Germany) and shipped on dry ice to the Sequencing and Genomics Facility of the Institute of Applied Biosciences, Center for Research and Technology Hellas, Thessaloniki, Greece for 16s rRNA analysis. Bacterial diversity was assessed by sequencing the V3-V4 regions of the 16S rRNA gene (∼460 bp) using the Illumina’s 16S Metagenomic Sequencing Library Preparation protocol. For the amplification of the V3-V4 region, gene-specific primers were selected based on Klindworth et al. 2013 (Klindworth et al. 2013), by adding Illumina overhang adapter nucleotide sequences at the 5′ end. Libraries were sequenced in a MiSeq platform using the MiSeq® reagent kit v3 (2×300 cycles) (Illumina Inc., San Diego, CA, USA).

### 2.5 Sequencing data processing

Sequencing reads were denoised into amplicon sequence variants (ASVs) using DADA2 (Callahan et al. 2016). Taxonomy was assigned to ASVs against the SILVA 132 16S rRNA database (Quast et al. 2013). Sequences classified as archaeal, chloroplastic or mitochondrial were removed. The final ASV table (3,027 ASVs in total) was imported into the phyloseq package in R for downstream analyses.

### 2.6 Statistical analysis

Continuous variables were summarized by mean and standard deviation, while categorical variables were presented by count and percent prevalence. To assess differences between the diagnosis groups, we used Welch’s t-test for the normally distributed data, Mann-Whitney U test for the non-normal data and Fisher’s exact test for categorical data. Statistical significance was set at a p-value <0.05.

Rarefied data (21,325 sequences per sample) were used for calculating alpha and beta diversity indices. Alpha diversity indices (richness and Shannon diversity index) were calculated using the phyloseq (v1.42) estimate_richness function and compared between groups using the Wilcoxon rank sum test. Beta diversity, based on Bray-Curtis dissimilarity, was analyzed using permutational multivariate analysis of variance (PERMANOVA, 999 permutations) from the vegan (v2.6-4) R package (“adonis2” function) (Oksanen J et al. 2022). Results were plotted by principal coordinate analysis (PCoA).

For differential abundance analysis, ASVs were aggregated at genus level and only genera with prevalence >0.1 (174 genera) were included. We applied Microbiome multivariable Associations with Linear model (MaAsLin2 v1.12.0) (Mallick et al. 2021) to detect differentially abundant genera, by fitting a zero-inflated negative binomial model in rarefied abundance data. Considering that the input data were rarefied, we turned off the default normalization and transformation methods implemented in MaAsLin2 function. Fixed effects included age, sex and MDS. P-values were adjusted for multiple testing using the false discovery rate (FDR). Genera with an adjusted p-value (q-value) lower than 10% were considered as differentially abundant genera.

To explore the discriminative performance of gut microbiota for amyloid-pathology and MCI, we regressed binary variables (amyloid-pathology or MCI) against the 174 identified genera. Due to the high dimensionality, sparsity and multicollinearity of microbiota data, we applied an elastic net regularized logistic regression on centered log-ratio (CLR) transformed counts (R 1996). This method can perform both feature selection and regularization during model training, and can often identify a small number of highly relevant features that capture the underlying patterns in the data. To evaluate the model performance, we obtained a 2×2 confusion matrix using the test dataset and calculated true positive, true negative, false positive, and false negative using predicted and observed classes. Then, we calculated and adopted three metrics for the model’s performance evaluation: accuracy, F1-score and area under the curve (AUC), using true positive rate and false positive rate (FPR) (Chicco and Jurman 2020; Metz 1978). Further information is provided in the **Supplemental Methods**.

To explore correlations of the levels of CSF biomarkers amyloid Aβ_42_, pTau and tTau with gut microbiome abundance data, we transformed counts with CLR (after applying a pseudocount of 0.0001) using *clr* function from the compositions R package (version 2.0-6). Spearman’s rank correlations were calculated using the *cor.test* function in R and considered statistically significant at *p*<0.05. To handle microbiome sparsity issues, we tested correlations only for genera harbored in at least 30% (≥15) of participants. The correlation heatmap was generated using the package ggcorrplot (v0.1.4.1). We used Phylogenetic Investigation of Communities by Reconstruction of Unobserved States 2 (PICRUSt2) (Douglas et al. 2020) to detect predicted functional differences in microbial communities across both comparisons (individuals with amyloid-pathology vs. amyloid- normal and CN vs. MCI). Predicted functionalities were further clustered in different pathway hierarchies (levels 1, 2, and 3) and compared across groups using multivariable logistic regression analysis with LASSO penalty and minimum lambda. All statistical comparisons and data visualization were performed with the R statistical programming language (v.4.2.1). All p values were 2-tailed (α = 0.05).

## 3. Results

### 3.1 Characteristics of the study population

We included 50 participants aged 64.3 (8.4), and 35 of them (70%) were female. **Table 1** summarizes the characteristics of the total sample and the diagnosis subgroups. Individuals with amyloid-pathology had lower amyloid values and were more likely to be carriers of *APOE* ε4, compared to amyloid-normal participants. Both groups were comparable in terms of sex, age, smoking status and MDS. When clinical diagnosis criteria were applied, 80% of overall participants (40 individuals) were classified as cognitively normal. As expected, age differed across clinical groups, as individuals with MCI were older. In regard to the levels of CSF biomarkers, amyloid was lower and pTau was higher in the MCI group, whereas tTau trended to being significantly different across groups. MCI and CN individuals were comparable in terms of sex, *APOE* ε4 positive status, smoking status and MDS. Distributions of CSF amyloid Aβ, pTau and tTau levels are presented in **Fig. S2**.

**Table 1.**
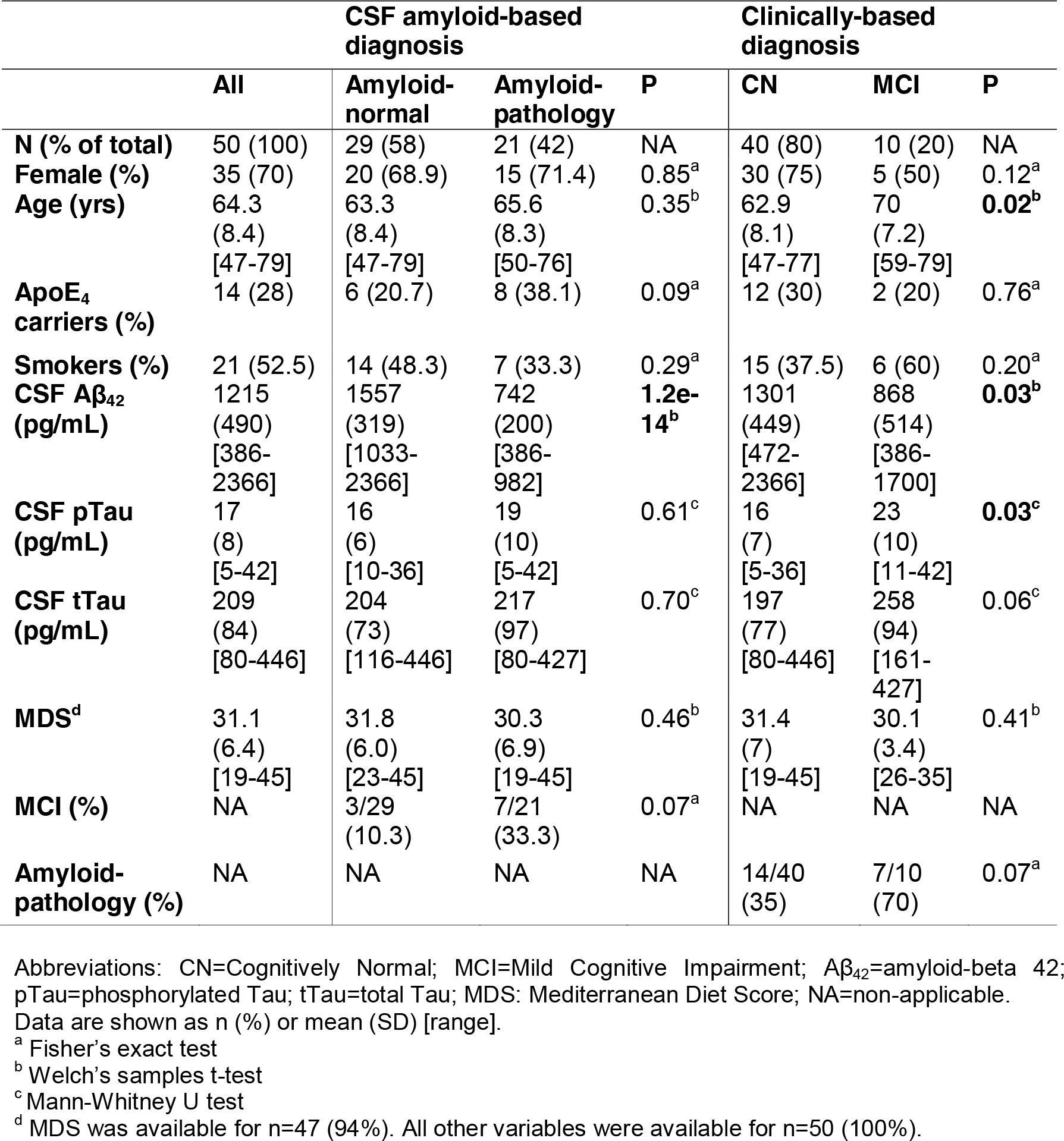
Descriptive characteristics of participants in groups according to diagnosis criteria CSF amyloid-based diagnosis criteria.

We profiled the gut microbiota through 16s rRNA amplicon sequencing and generated a total of 1,936,604 high-quality reads with an average of 38,732 reads per sample. Among all 50 participants included in this analysis, the dominant phyla were Firmicutes and Bacteroidetes, which made up 67% and 25% of the total abundance, respectively (**Fig. S3, S4**). The Firmicutes/Bacteroidetes ratio did not differ either between CSF amyloid-based diagnosis groups (mean [SD], amyloid-normal 2.84 [0.95]; amyloid-pathology 3.03 [1.41], P=0.60) or between clinically-based diagnosis groups (mean [SD], CN 2.83 [0.96]; MCI 3.29 [1.75], P=0.44). The predominant bacterial families, overall, were *Lachnospiraceae* (∼30.8%), *Ruminococcaceae* (∼24%), and *Bacteroidaceae* (∼15.5%) (**Fig. S3, S4**). The most abundant five genera across all individuals were *Bacteroides* (∼15.5%), *Faecalibacterium* (∼8.5%), *Blautia* (∼5.8%), *Eubacterium hallii group* (∼4.3%) and *Ruminococcus 2* (∼3.5%) (**Fig. S3, S4**), however no differences were found across the diagnosis groups. Similarly, no differences in alpha diversity, measured with richness and Shannon diversity index, and in beta diversity (PCoA, Bray-Curtis) were found in either comparison type (**Fig. 1**, **Table S1**).

**Figure 1.**
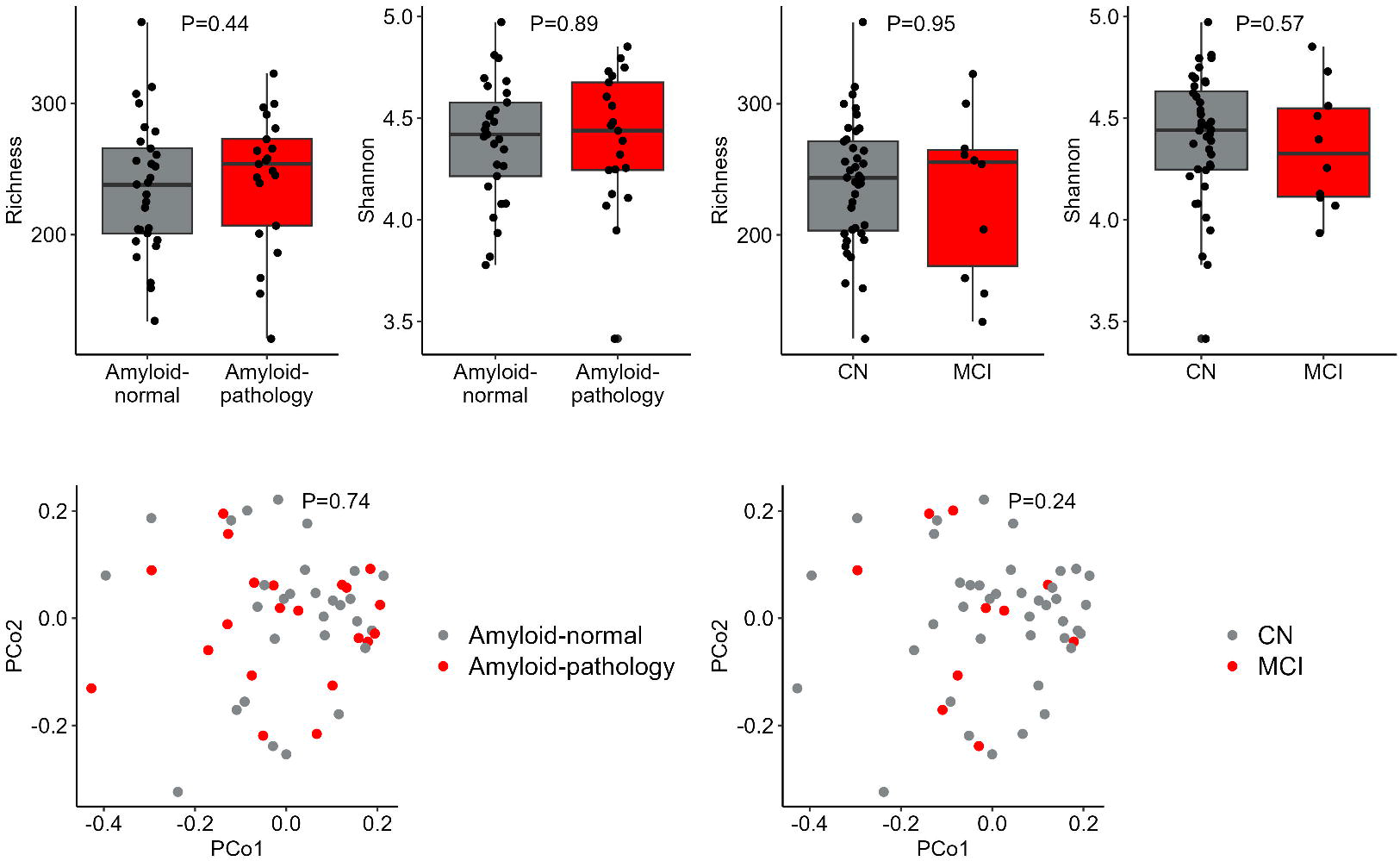
Alpha and beta diversity are similar among groups, according to diagnosis status. In the upper panel, boxplots of alpha diversity measures (richness and Shannon diversity) are shown. P-values are from Wilcoxon rank sum test. In the bottom panel, beta diversity is presented by PCoA (measured by Bray-Curtis distance). P-values are from PERMANOVA test (999 permutations), adjusted for age, sex and Mediterranean diet score. Abbreviations: PCoA=Principal coordinate analysis; CN=cognitively normal; MCI=Mild cognitive impairment.

### 3.2 Associations of microbiota with amyloid pathology and MCI status

Differential abundance (DA) analysis revealed that gut microbiota of individuals with amyloid-pathology showed significantly altered abundances of 21 genera (FDR<0.10) relative to the amyloid-normal group, with 10 genera more abundant and 11 genera less abundant in amyloid-pathology group (**Table S2**). Genera detected in at least 30% of participants and most associated with amyloid-pathology status by magnitude of their model coefficients included *Oxalobacter* (coefficient=1.06, standard error=0.28, q-value=0.003), *Coprobacter* (coefficient=0.81, standard error=0.33, q-value=0.099) and *Marvinbryantia* (coefficient=0.71, standard error=0.26, q-value=0.049). *Enterococcus* (coefficient=-2.55, standard error=0.95, q-value=0.053) and *Enterobacter* (coefficient=-1.24, standard error=0.38, q-value=0.013) were found to be the top two genera associated with amyloid-normal status (**Fig. 2**). When individuals with MCI were compared to CN individuals, we found 27 differentially abundant genera (FDR<0.10), of which 19 were more abundant in the MCI group (**Table S3**). The top three genera detected in at least 30% of participants and most associated with MCI status by magnitude of their model coefficients included *Escherichia-Shigella* (coefficient=2.60, standard error=1.03, q-value=0.072), *Family XIII AD3011 group* (coefficient=1.61, standard error=0.46, q-value=0.007) and *Enterobacter* (coefficient=1.39, standard error=0.43, q-value=0.014). *Dialister* (coefficient=-1.92, standard error=0.51, q-value=0.003) was the genus most associated with CN status (**Fig. 2**).

**Figure 2.**
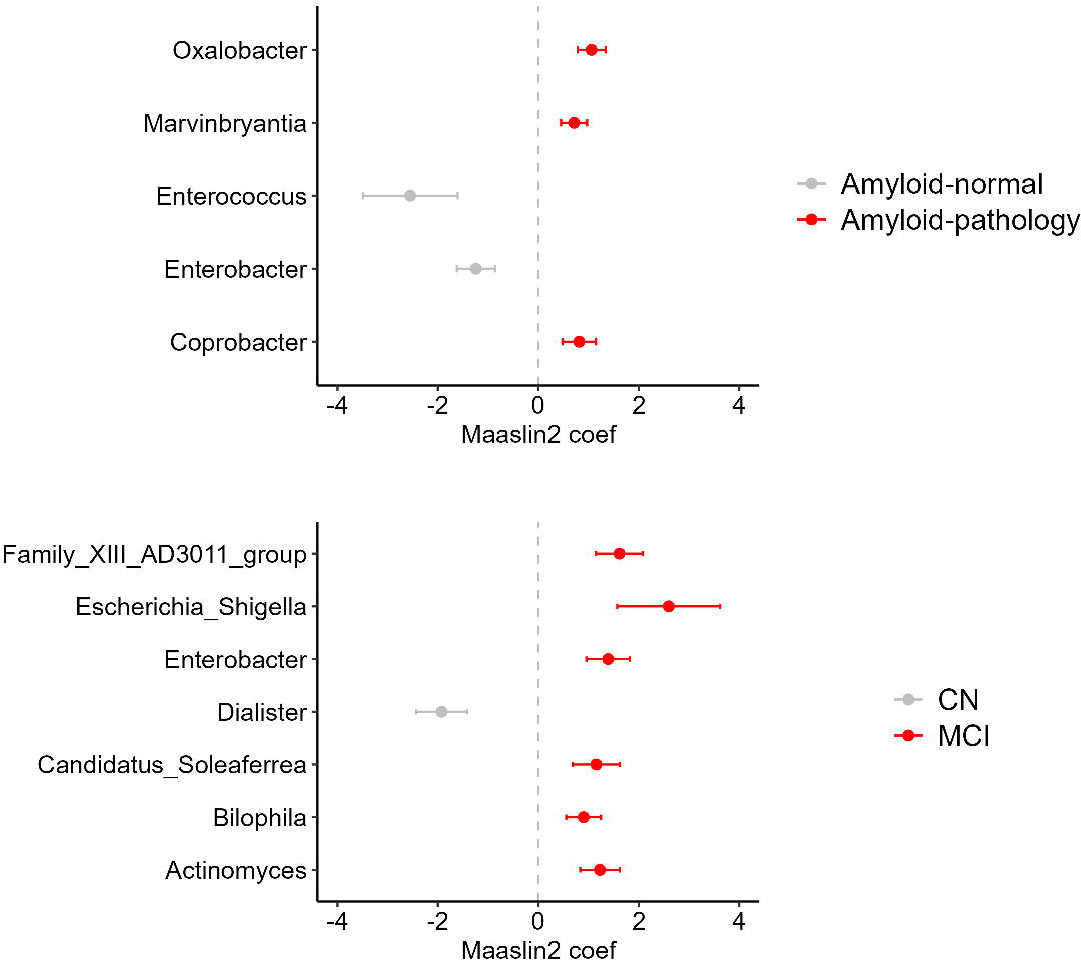
Specific gut microbial genera are associated with amyloid-pathology or MCI. Differentially abundant genera with a minimum prevalence of 30% and FDR adjusted p- value<0.10 identified through the statistical MaAsLin2 R package are shown. The x axis shows the effect size represented by MaAsLin2 coefficient. Taxa coefficients are from negative binomial regression models (as implemented in MaAsLin2) that additionally included age, sex and Mediterranean diet score as predictors. Positive number means enrichment in amyloid-pathology or MCI group (red color), while negative value means enrichment in the amyloid-normal or CN group (grey color)). All regression model results are presented in **Tables S2** and **S3**. Abbreviations: CN=Cognitively normal; MCI=Mild cognitive impairment.

### 3.3 Common and distinct microbiota between amyloid pathology and MCI status

Of the DA genera identified at both comparisons, six genera (after removal of unclassified genera) were shared (**Fig. S5**), of which only *Enterobacter* showed opposite direction of effect, i.e. lower abundance in individuals with amyloid-pathology and higher abundance in individuals with MCI (**Fig. 2**). The remaining 28 DA genera were found to uniquely differentiate amyloid-based or clinically-based diagnosis groups, with their direction of effect reported in **Fig. S5**. Based on the MaAsLin2 association p-values (**Tables S2, S3**) and genera prevalence (detected in at least 30% of individuals), *Oxalobacter, Coprobacter, Marvinbryantia* and *Enterococcus* may differentiate only amyloid-based pathology groups, through lack of statistically significant association in the clinically-based comparison. Similarly, genera *Bilophila, Actinomyces* and *Escherichia-Shigella* may differentiate only clinically-based diagnosis groups, through lack of statistically significant association in the amyloid-based comparison.

### 3.4 Discriminative capacity of microbiota for amyloid pathology and MCI status

The elastic net analyses comparing participants with or without CSF amyloid-pathology identified two microbial genera discriminating the amyloid status, with *Coprobacter* over- represented and *Oscillibacter* under-represented in pathology (**Fig. 3A**, **Table S4**). The overall predictive accuracy of the classification model (AUC) was 0.778 (95%CI=0.774- 0.783, sensitivity=0.992, specificity=0.574). Our model had moderate diagnostic accuracy based on the confusion matrix, the F1 score (0.781) and the overall prediction accuracy (0.752). These microbial genera remained in the model after adjusting for age, sex and MDS (**Table S4)**. For MCI status, the identified signature achieved a greater AUC of 0.928 (95%CI= 0.926-0.931, sensitivity=0.789, specificity=0.703) with *Escherichia-Shigella* and *Lachnospiraceae UCG-004* over-presented, and *GCA-900066575* under-represented (**Fig. 3B**, **Table S5)**. Based on the confusion matrix, the F1 score (0.896) and overall accuracy (0.850), our model has high diagnostic accuracy. Sensitivity analysis did not affect our unadjusted model results, however two new genera (*Oxalobacter;* coefficient=0.035; 95% CI: 0.020, 0.049 and *Enterobacter*; coefficient=0.052, 95%CI: 0.037, 0.067) were added as important features (**Table S5**). Of note, *Escherichia-Shigella and Enterobacter* were defined as DA genera from MaAsLin2 analysis and in the same direction (i.e. higher in the MCI group) (**Fig. 2**).

**Figure 3.**
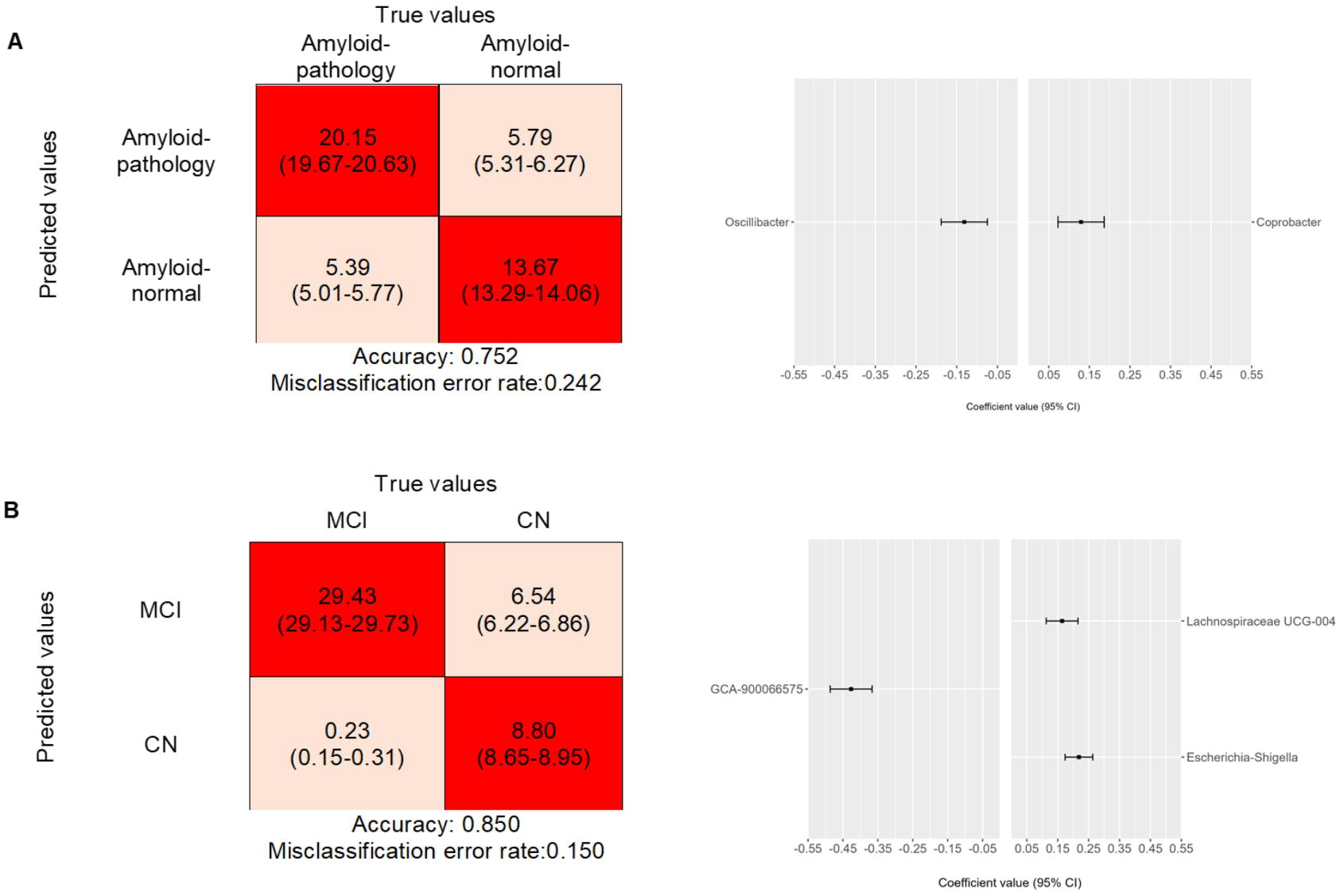
Gut microbiota have a fairly good discriminative performance for amyloid pathology and MCI. Confusion matrix obtained from the regularized logistic regression model and genera ranked from the highest to the lowest elastic net positive and negative regression coefficients are shown for **A)** CSF amyloid-based and **B)** clinically-based diagnosis. For a given confusion matrix, the x-axis shows each of the two targets while the y- axis shows each of the two predicted labels. The upper left and lower right squares display the values of correct classification, while the upper right and lower left squares display the values of misclassification.

### 3.5 Correlation of microbiota with CSF biomarkers for AD

We calculated Spearman’s rank correlations between the CLR-transformed abundance of genera identified as differentially abundant or as important features through elastic net models and the three measured CSF biomarkers (amyloid, pTau and tTau) (**Fig. 4**). The MCI-abundant *Escherichia-Shigella* (rho=-0.33, P=0.010) and the amyloid-pathology abundant *Marvinbryantia* (rho=-0.27, P=0.026) were significantly negatively correlated with amyloid levels, while *Oscillibacter*, a genus with lower abundance in the amyloid-pathology group based on the elastic net regression model, was significantly negatively correlated with pTau levels (rho=-0.22, P=0.046) (**Fig. S6**). None correlation survived FDR<5% correction. No genera were significantly correlated with tTau levels.

**Figure 4.**
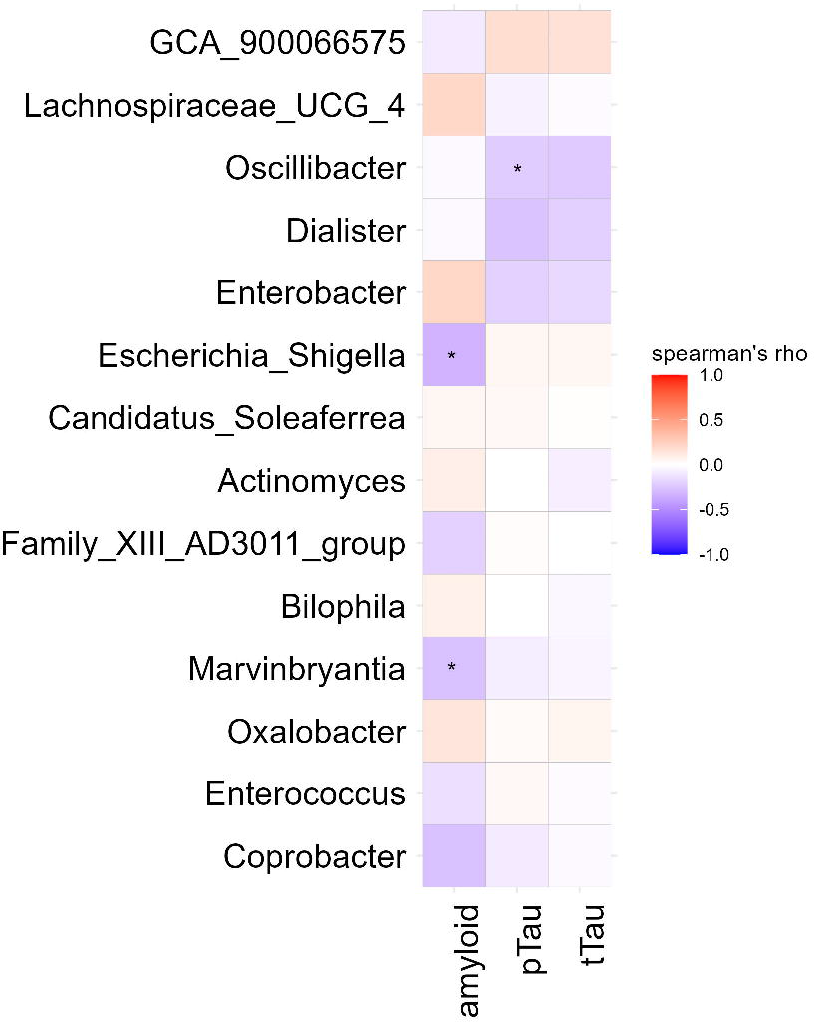
Heatmap of spearman correlation analysis between genera of interest and levels of CSF biomarkers. The genera of interest are the differentially abundant genera, as detected from MaAsLin2 (in ≥30% of individuals), and genera selected from elastic net regression models (including those from sensitivity analysis). CLR-transformed counts were used for the correlation analysis. The rho values are represented with gradient colors, where blue and red cells indicate negative and positive correlations, respectively. Significant correlations are highlighted with an asterisk (* *p*<0.05, unadjusted).

### 3.6 Prediction of metagenomes

A multivariable logistic regression analysis using LASSO penalty with minimum lambda was applied to detect significant KEGG pathways that differentiate diagnosis groups. When individuals with amyloid-pathology were compared to amyloid-normal individuals, we found a decrease in the pathway (i.e. gene content) related to environmental adaptation (2^nd^ level) in the amyloid-pathology group. When individuals with MCI were compared to CN individuals, in level 3, pathways related to endocytosis, polycyclic aromatic hydrocarbon degradation and stilbenoid, diarylheptanoid and gingerol biosynthesis (3^rd^ level) were increased in the MCI group (**Fig. S7**).

## 4. Discussion

We profiled the gut microbiota in both early preclinical stage of AD (as reflected from amyloid pathology) and MCI in a well-characterized cohort of nondemented individuals. Our analysis demonstrated that distinct microbial alterations may be present in each stage of the disease continuum. Furthermore, we developed microbiota signatures that have good discriminative performance for amyloid pathology and MCI status and showed that levels of differentially abundant microbiota were correlated with CSF biomarkers of AD pathology. Our findings add to the emerging literature highlighting the potential utility of gut microbiota as complementary early prognostic markers for AD.

Our DA analysis revealed higher levels of *Oxalobacter* in individuals with amyloid pathology, a finding that has not been previously reported in other microbiome studies exploring preclinical stage of AD (Verhaar et al. 2021; Jung et al. 2022; Ferreiro et al. 2023). We also found over-representation of *Oxalobacter* in MCI individuals through elastic net analysis, however results on this genus in relation to MCI are inconsistent across different studies (Chuang 2022; Fan et al. 2023). *Oxalobacter* has been shown to metabolize oxalate (Daniel et al. 2021), which in turn may be involved in the formation of amyloid aggregates in the entorhinal cortex of AD specimens, however its role in cognitive decline remains unclear (Heller, Coffman, and Jarvis 2020). Oxalate has been also shown to promote inflammation and systemic oxidation through inducing reactive oxygen species (Liu et al. 2021). Another finding of an amyloid-pathology abundant genus is *Marvinbryantia*. We found an opposite direction of effect compared to the findings of a study of individuals from the Netherlands (Verhaar et al. 2021), which revealed an association of lower levels of *Marvinbryantia* with higher odds of amyloid positivity. This discrepancy may be due to the 3-fold smaller sample size of our study and subsequent lack of power or due to other factors including methodological differences, confounding effects of factors influencing gut microbiome composition (e.g. living environment, lifestyle) or differences in analysis pipelines. In our study, *Marvinbryantia* was the only genus associated with amyloid-pathology, and also correlated with lower CSF amyloid levels. To our knowledge, there are no other studies reporting an association of this microbe and AD biomarkers. *Marvinbryantia* belongs to the *Lachnospiraceae* family, known for the production of SCFAs such as butyrate, which regulates gut permeability and stimulates the inflammatory response (Siddiqui and Cresci 2021). We also report an association of lower abundance of *Enterococcus* with amyloid-pathology, in line with a previous study (Jung et al. 2022). *Enterococcus*, a lactic-acid producer, has been shown to have anti-inflammatory properties (Carasi et al. 2017) and, thus, lower levels of *Enterococcus* may accelerate brain amyloid-β deposition through increased inflammation (Kinney et al. 2018).

We highlight *Escherichia-Shigella* among the top genera associated with MCI status. This pathogenic microbe was found at higher levels in individuals with MCI, and also correlated with lower amyloid levels. Consistent to our findings, a study of individuals from Turkey also reported an association of this genus with MCI (Yildirim et al. 2022). *Escherichia-Shigella* belongs to the *Enterobacteriaceae* family, which can produce endotoxins (e.g. lipopolysaccharide) that are released into the blood circulation and thus induce systemic inflammation. The abundance of *Escherichia-Shigella* has been previously positively correlated with levels of proinflammatory cytokines such as IL-1β and NLRP3 (Cattaneo et al. 2017). On the other hand, *Dialister* was the top genus associated with CN status, in accordance with previous study (Vogt et al. 2017), implying a protective role against development or progression of AD pathology.

Our analyses allowed us to identify microbiome features that specifically differentiate amyloid-pathology and clinically-based diagnosis groups. We revealed that the microbiota alterations that we observed in individuals with amyloid pathology are only partly found in individuals with MCI. Our findings extent recent literature reporting stage-specific microbial alterations in MCI and AD (Troci et al. 2024) by revealing distinct microbial communities in the early preclinical stage of CSF amyloid-based pathology, where clinical symptoms are absent. Among the overlapping microbiota, notably, only *Enterobacter* showed opposite directionality of effects i.e. lower in amyloid pathology and higher in MCI. *Enterobacter* has been previously shown to increase steadily from healthy controls to MCI and dementia stage (Chen et al. 2023; Liu, Wu, et al. 2019), consists of pro-inflammatory species and correlates with brain structural signatures in regions related to memory, emotional processing and cognition (Tsai et al. 2022). Different abundance levels of *Enterobacter* between early preclinical stages and MCI may indicate a progressive change of gut microbiota composition during AD continuum, potentially opening future opportunities for gut-directed interventions that could interdict progression to clinical AD. Studies in larger cohorts are needed to validate our results.

The use of gut microbiota as predictive markers for AD has shown promise (Ferreiro et al. 2023; Verhaar et al. 2021). There is a clear attraction in the development of a gut microbiota-based summary signature that can be used in both early preclinical stage of AD and MCI for risk stratification. Here, we demonstrated a fairly good discriminative capacity of gut microbiota for both amyloid pathology and MCI. Two genera, *Oscillibacter* and *Coprobacter,* were found as important features for amyloid-pathology status. *Oscillibacter* has been previously associated with preclinical AD and reported as an important predictor (Ferreiro et al. 2023), however, the direction of effect was opposite (higher abundance associates with preclinical AD) compared to our study. This supports the need to expand microbiome studies in different populations and highlights the importance of host and environmental context, disease stage and strain-specific effects. *Oscillibacter spp.* are known to correlate to decreased colonic epithelial integrity (Lam et al. 2012), while *Oscillibacter sp. 57_20* has been associated with positive indicators of cardiometabolic health in a recent microbiome-wide association study (Asnicar et al. 2021). Moreover, *Oscillibacter* showed significant alterations from MCI to AD but not from normal cognition to MCI (Zhu et al. 2022). Notably, in our study, lower abundance of *Oscillibacter* correlated with higher pTau levels but not with total tau, which is a marker of neurodegeneration (Craig-Schapiro, Fagan, and Holtzman 2009), in line with previous report (Ferreiro et al. 2023). *Coprobacter*, also found as amyloid-pathology abundant in our study, is a SCFA-producer, that has been found in higher levels in amnestic MCI (Duan et al. 2021) and in children with autism spectrum disorder (Liu, Li, et al. 2019). The increase of *Coprobacter* could act as a compensation mechanism to maintain high levels of SCFAs during the initial phases of the disease, however this warrants further elucidation. For MCI status, a signature of three genera, namely *Escherichia-Shigella*, *Lachnospiraceae-UCG-004* and *GCA-900066575*, demonstrated a high level of discriminative performance. *Escherichia-Shigella* was also found as DA genus associated with MCI. Our results add to current knowledge that gut microbiome may be helpful for detecting or screening early stages of the disease, however larger studies are needed to confirm our findings.

Our research has several strengths. We revealed distinct gut microbiota profiles associated with amyloid-β pathology, and with MCI. The availability of both CSF samples and clinical data from early disease stages allowed us to perform simultaneous comparison of participants based on both clinical evaluation and the assessment of early biomarkers, making our study novel. Second, the study population was derived from the very well-characterized ALBION cohort that has been recruited in a specialist clinic of a tertiary university hospital. In addition, clinical diagnosis was established by a panel of experts after an extensive standardized neuropsychological assessment. Third, we used advanced modeling analysis techniques to profile gut microbiota of individuals from different stages of AD continuum. Microbiota composition was determined by 16s rRNA amplicon sequencing, which is a widely used sequencing method. Machine learning model helped us identify gut microbiota features with good discriminative capacity for amyloid pathology and MCI status. Finally, our study sample includes adults across a broad age range, in contrast to past literature that has primarily focused in older (mean age >73yrs) adult populations (Jung et al. 2022; Ferreiro et al. 2023). However, there are important limitations that need to be considered. First, due to the cross-sectional study design, we cannot infer causal associations of gut microbiota with brain Aβ deposition. However, longitudinal fecal sampling and biomarkers assessments within ALBION study are ongoing (so far up to five years for a few participants), enabling us to identify gut microbiome changes that associate with progression of individuals to MCI or symptomatic AD. Second, although we adjusted for relevant confounders as age, sex and Mediterranean Diet Score (as surrogate for dietary intake), we cannot rule out unmeasured and residual confounding effects. Other confounding variables such as obesity, physical inactivity, hypertension and hypercholesterolemia, which have been shown to be important dementia risk factors(Bransby et al. 2024), also need to be further controlled. Furthermore, we did not consider the time interval between CSF assessments and fecal sampling, raising the possibility that biomarker values used in the analysis may differ from true biomarker levels at the time of fecal sampling. Third, our study interrogated gut microbiota profiling through 16s rRNA amplicon sequencing. As a result, we cannot define species, but also, functional pathways, precisely. We revealed pathways involved in amyloid-β dynamics and pathology (e.g. environmental adaptation, (Kress et al. 2018)) or brain deficits in the onset of MCI (e.g. endocytosis, (Zadka et al. 2024)), however a cautious interpretation of PICRUSt2 findings is warranted. Finally, the sample size was relatively small compared to other studies (Ferreiro et al. 2023; Verhaar et al. 2021), thus making it difficult to reveal subtle differences between groups. However, we were able to provide an overview of gut microbiota profiles of both diagnosis groups defining microbiome differences at a stringent FDR threshold (5%).

## 5. Conclusions

In summary, our findings suggest that distinct gut microbiome features may associate with early preclinical stage for AD as reflected by CSF amyloid-β pathology, and with MCI. We further demonstrate a discriminative capacity of gut microbiota for amyloid pathology and MCI status. Overall, our findings may contribute to the discovery of diagnostic markers potentially be used from the clinicians as early detection markers and potential therapeutic targets for AD. However, additional research is needed to validate our results in broader preclinical AD cohorts, assess causal effects and investigate whether these associations extend to individuals with AD symptoms.

## CRediT authorship contribution statement

**KR:** Conceptualization, Data curation, Formal analysis, Investigation, Methodology, Resources, Visualization, Writing - Review & Editing, Writing - Original Draft. **EM:** Investigation, Methodology, Resources. **EN:** Investigation, Methodology, Resources. **MP:** Investigation, Methodology. **MA:** Investigation, Methodology. **CE:** Formal analysis Methodology. **NNF:** Data Curation, Formal analysis, Methodology, Visualization, Writing – Review & Editing. **MB:** Methodology. **ASD:** Conceptualization, Resources, Writing - Review & Editing. **CP:** Conceptualization, Data Curation, Formal analysis, Visualization, Writing – Review & Editing. **MY:** Conceptualization. **AA:** Conceptualization, Resources, Supervision**. NS:** Conceptualization, Project administration, Supervision, Writing - Review & Editing.

## Data availability statement

All data used in the analyses of this study are available within the manuscript and its supplemental information files. The raw sequencing data generated from this study have been deposited in NCBI SRA (https://www.ncbi.nlm.nih.gov/sra) under the accession number PRJNA1066101.

## Submission declaration and verification

We declare that this manuscript has not been published previously nor being considered for publication elsewhere. All authors have given their approval for submission of the manuscript to be considered for publication in *Neurobiology of Aging*. If accepted, it will not be published elsewhere in the same form, in English or in any other language, including electronically without the written consent of the copyright-holder.

## Acknowledgements

We thank all the members of Aiginition Hospital of Athens for their contribution. We also sincerely thank the participants for their participation in this study.

## Footnotes

1 Abbreviations: AD, Alzheimer’s disease; Aβ_42_: amyloid-beta 42; MCI, Mild cognitive impairment; CSF, Cerebrospinal fluid; rRNA: ribosomal RNA; pTau: phosphorylated tau; tTau: total tau; APOE: Apolipoprotein E; MDS: Mediterranean Diet Score; ASV: Amplicon sequence variant; PERMANOVA, permutational multivariate analysis of variance; PCoA: Principal coordinate analysis; MaAsLin2, Microbiome multivariable Associations with Linear model; FDR, False discovery ratio; CLR, Centered log ratio; AUC, Area under the curve; PICRUSt2: Phylogenetic Investigation of Communities by Reconstruction of Unobserved States 2.

