## Supplemental Methods for "Distinct gut microbiota profiles may characterize amyloid beta pathology and mild cognitive impairment"

**Genera associated with classification based on clinical diagnosis.**

To deal with data imbalance (10 MCI and 40 cognitively normal) we used the Synthetic Minority Over-sampling Technique (SMOTE), a synthetic sampling method suggested for microbiome research. Through this approach new samples were synthesized in silico based on existing minority class samples and added to the training set. After applying it we had augmented the MCI cases to 45, which we used for estimating optimum alpha and lambda parameters. The regression coefficients were obtained using the original 10 MCI cases, applying weighted elastic net regression to correct for data imbalance. To test the performance of the signature we used the 10 MCI cases.

To identify a gut microbiota signature for MCI, we regressed diagnosis status (MCI vs cognitively normal) on the 174 genera using a logistic regression with elastic net penalty. To reduce overfitting, the data was divided into training set (90%) and test set (10%). In the training set, we evaluated the alpha parameter from 0.1 to 1 in ~0.05 increments (avoiding L2 norm), and the tuning parameter (lambda), using a leave-one-out cross-validation (given the small sample size) approach in a 30-iteration loop. The combination of alpha and lambda was chosen based on a high Area Under the ROC Curve (AUC) in the test set. We found that alpha = 0.70 and lambda = 0.173050841 gave the highest performance in the test set. We applied the selected alpha and lambda values to each penalized regression for every training (90% of our primary dataset) in a 10-iteration loop. The coefficients obtained from 10 iterations in the elastic net were applied to the selected genera as weights (positive or negative) to estimate the signature of MCI as the weighted sum. For each genus, we calculated the mean coefficient and the 95% CI.

To address potential confounding effects of age, sex, MDS we added them in the elastic net model as covariates (unpenalized).

**Genera associated with classification based on amyloid-pathology.**

We applied the same rationale as described earlier, but to reduce overfitting, the data was divided into training set (70%) and test set (30%). Also, we didn’t have to deal with data imbalance since the number of amyloid-pathology cases were 21 and amyloid-normal individuals were 29. We found that alpha = 1.00 and lambda = 0.12943823 gave the highest performance in the test set. To select genera, we ran 10 iterations and selected those genera that appeared 5 times. The coefficients obtained from 10 iterations in the elastic net (figure) were applied to the selected genera as weights (positive or negative) to estimate the signature of pathology as the weighted sum. We also performed sensitivity analysis adjusting the elastic net model for age, sex, MDS (unpenalized).
