## Supplementary Figures for "Distinct gut microbiota profiles may characterize amyloid beta pathology and mild cognitive impairment"

**Figure S1.** Study flowchart.

**Figure S2.** Distribution of the levels of CSF biomarkers (pg/ml) by diagnosis criteria.

**Figure S3.** Compositional plots of relative abundance among the amyloid-based diagnosis groups at phylum, family and genus level.

**Figure S4.** Compositional plots of relative abundance among the clinically-based diagnosis groups at phylum, family and genus level.

**Figure S5.** Shared and distinct gut microbiota across the two diagnosis-based comparisons.

**Figure S6.** Spearman’s rank correlation plots bewteen gut microbiota abundance and CSF AD biomarkers.

**Figure S7.** Functional pathways predicted by Picrust2 that differentiate diagnosis groups.

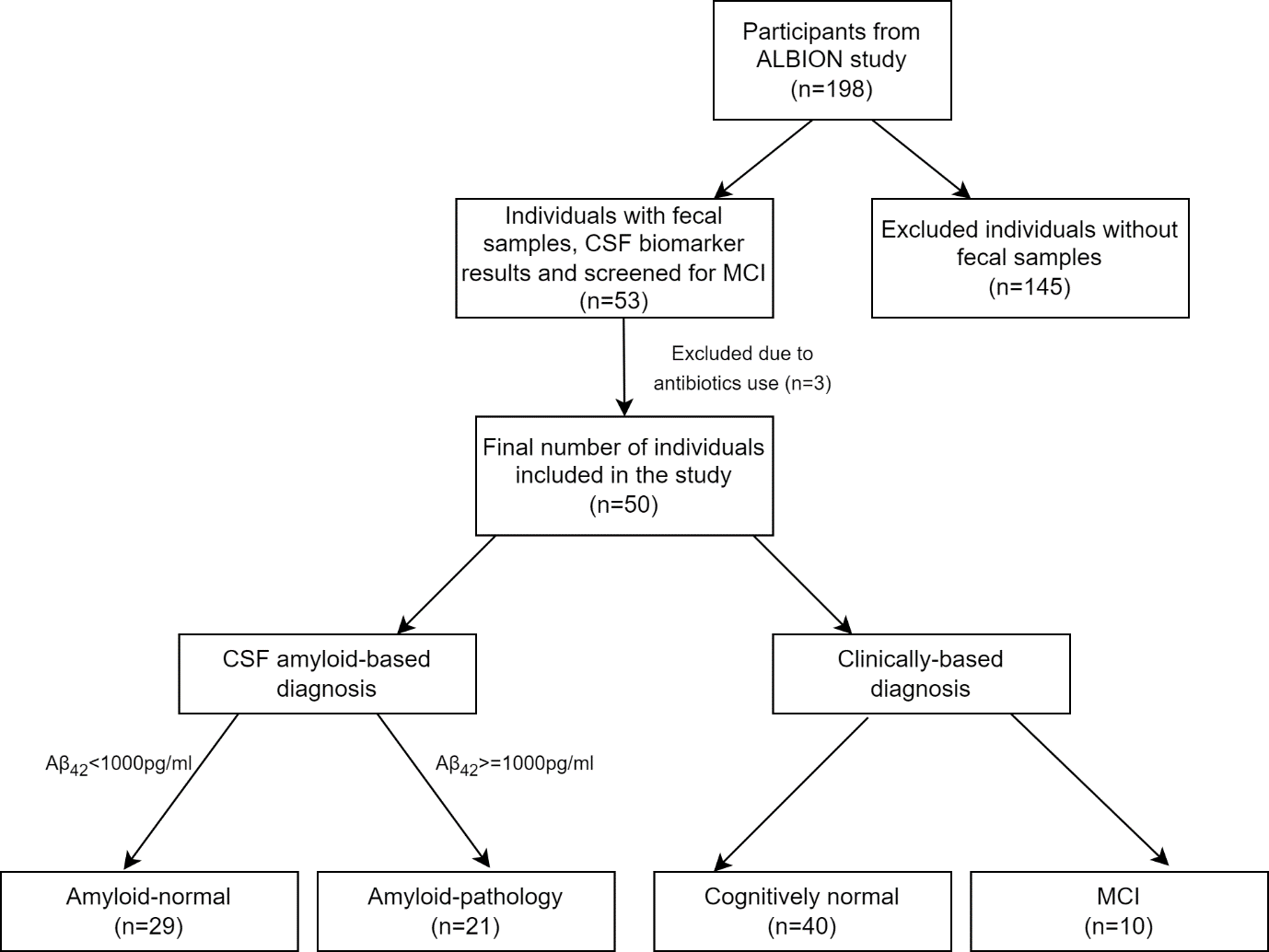

**Figure S1.** **Study flowchart.** Flowchart of the number of individuals from the ALBION (Aiginition Longitudinal Biomarker Investigation of Neurodegeneration) cohort invited for fecal sample collection, included in the analyses and classified based on diagnosis criteria. Aβ: amyloid-β. MCI: mild cognitive impairment.

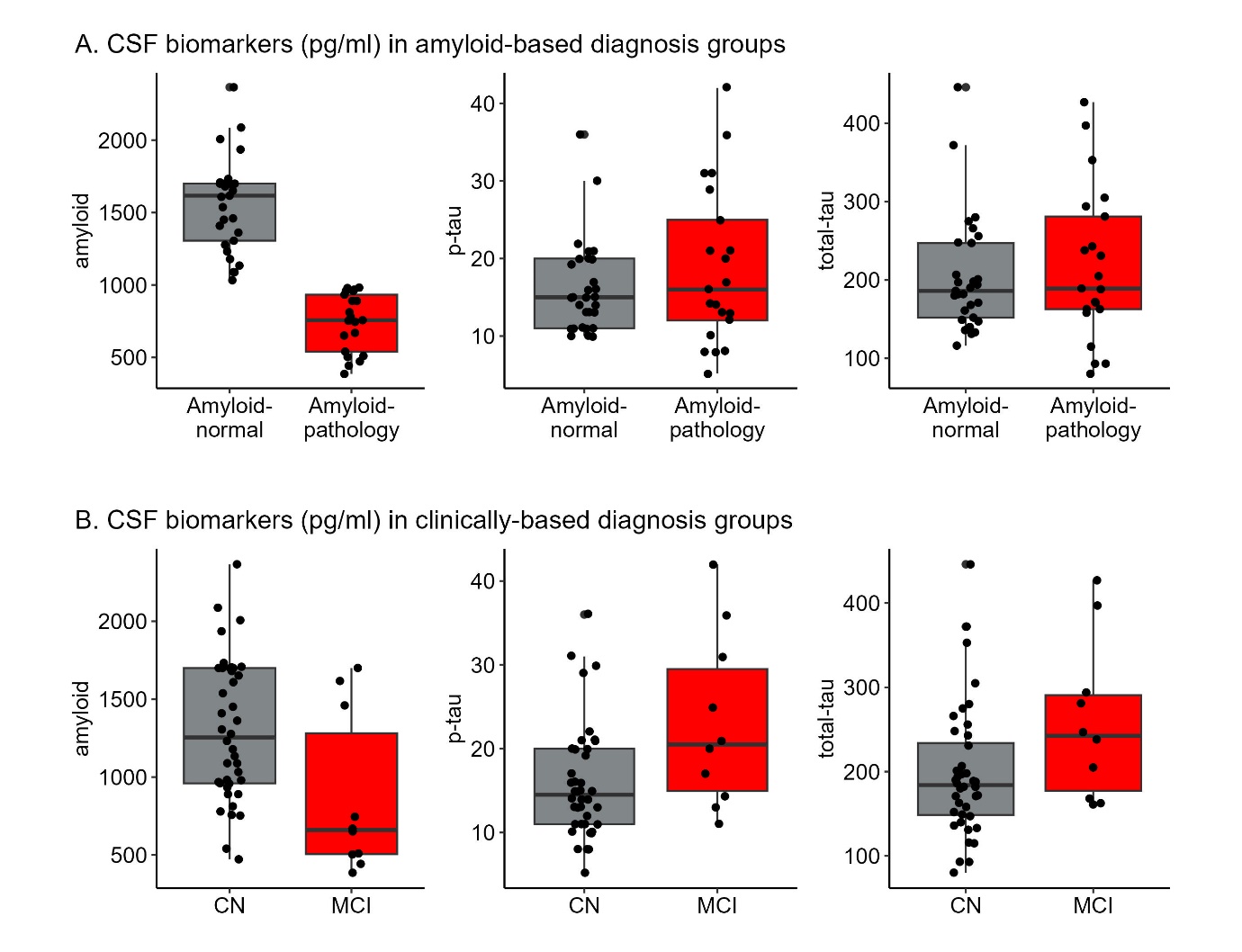

**Figure S2. Distribution of the levels of CSF biomarkers (pg/ml) by diagnosis criteria.** CN: Cognitively Normal. MCI: Mild Cognitive Impairment.

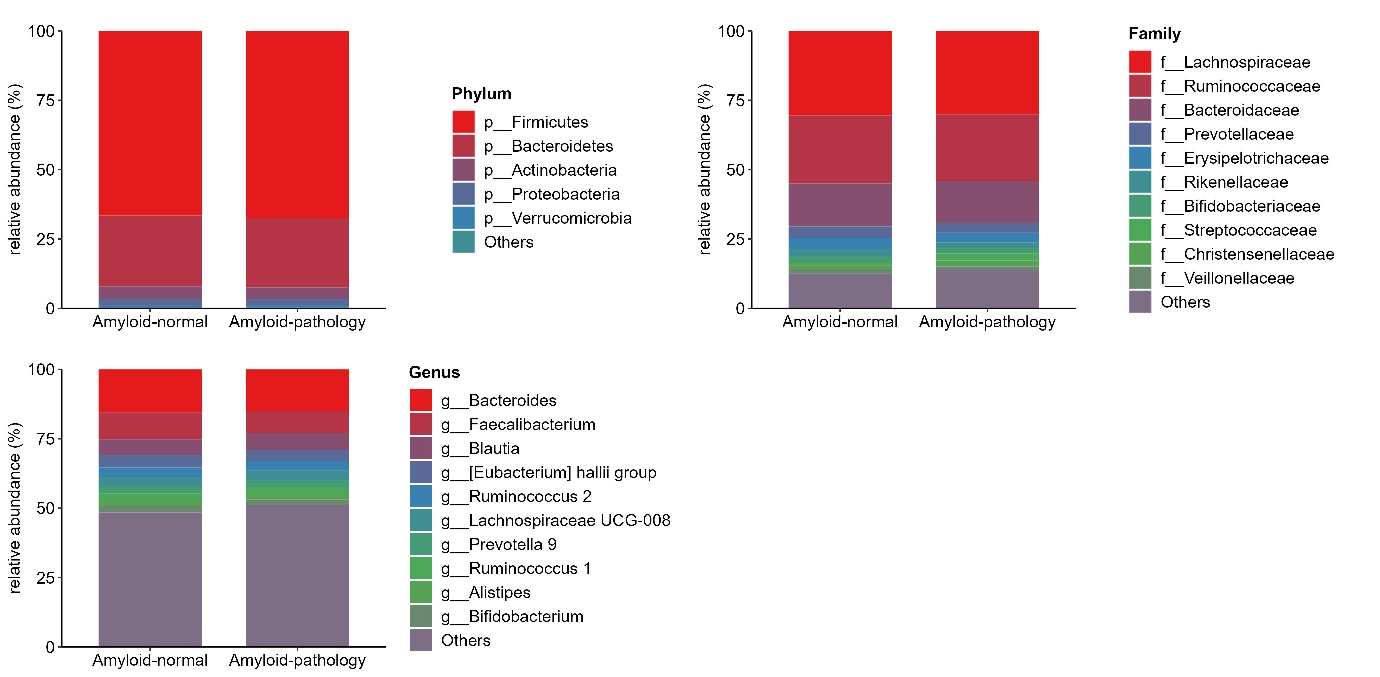

**Figure S3. Compositional plots of relative abundance among the amyloid-based diagnosis groups at phylum, family and genus level.**

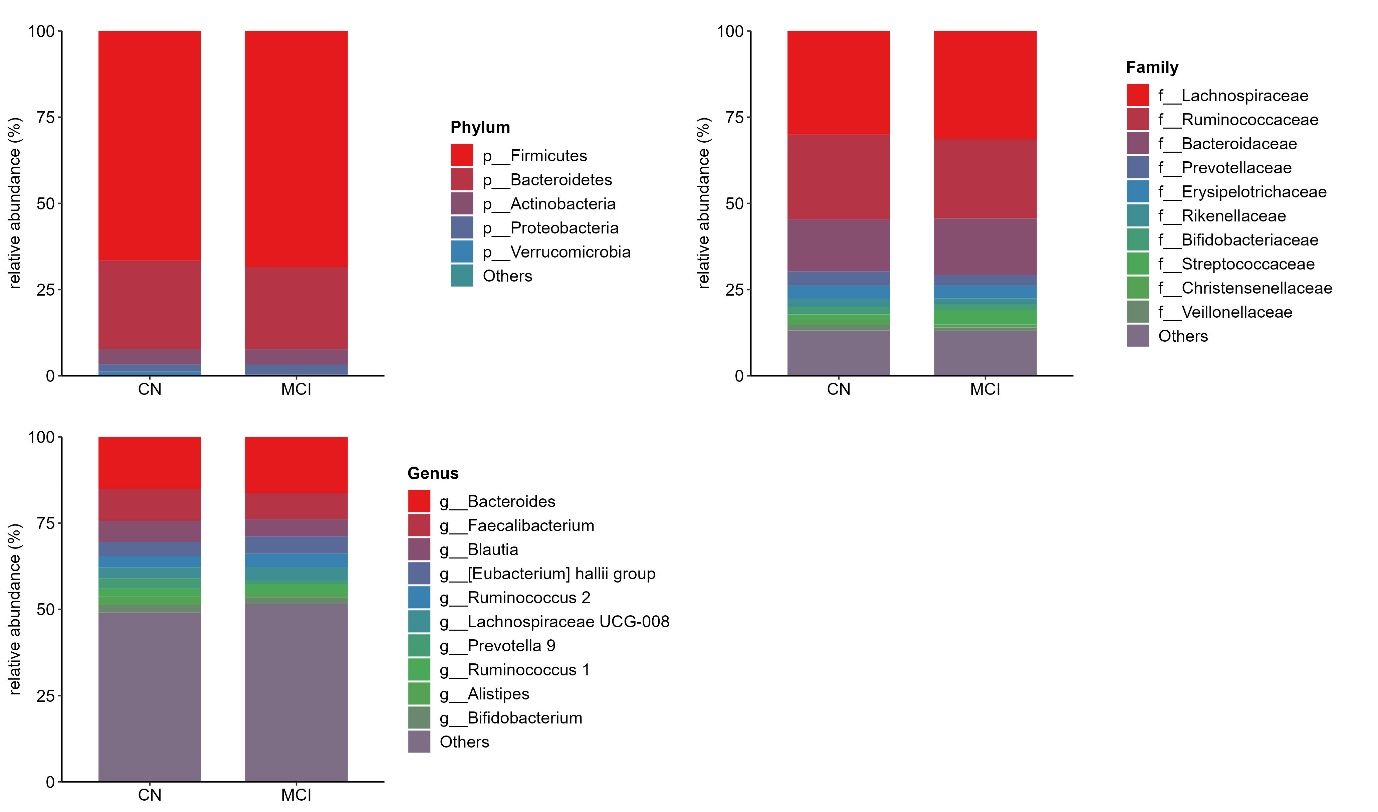

**Figure S4. Compositional plots of relative abundance among the clinically-based diagnosis groups at phylum, family and genus level.**

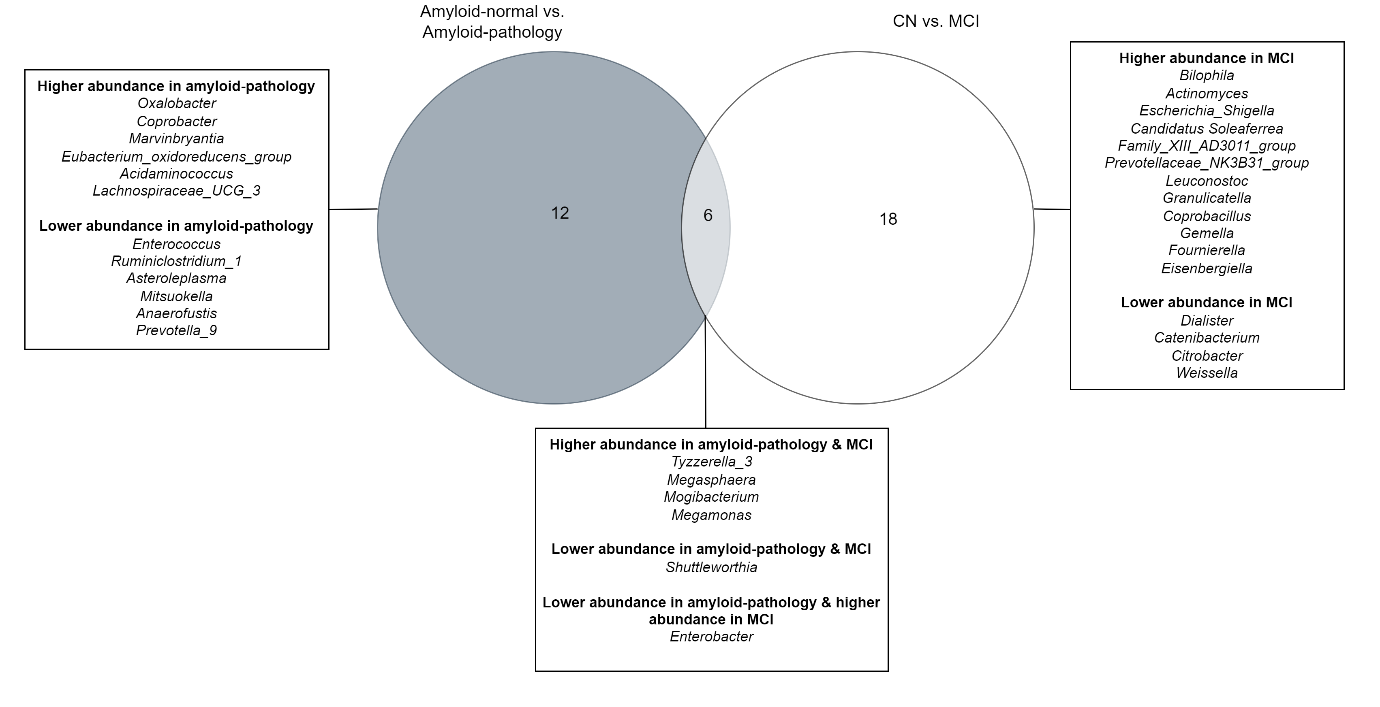

**Figure S5. Shared and distinct gut microbiota across the two diagnosis-based comparisons.** Venn diagram depicts overlapping MaAsLin2-derived DA genera across both types of comparison (amyloid-normal vs. amyloid-pathology and CN vs. MCI). Direction of effect is reflected by the higher or lower levels of abundance in amyloid-pathology or MCI group. DA genera that were assigned as unclassified are not shown. Abbreviations: CN=Cognitively normal; MCI=Mild cognitive impairment.

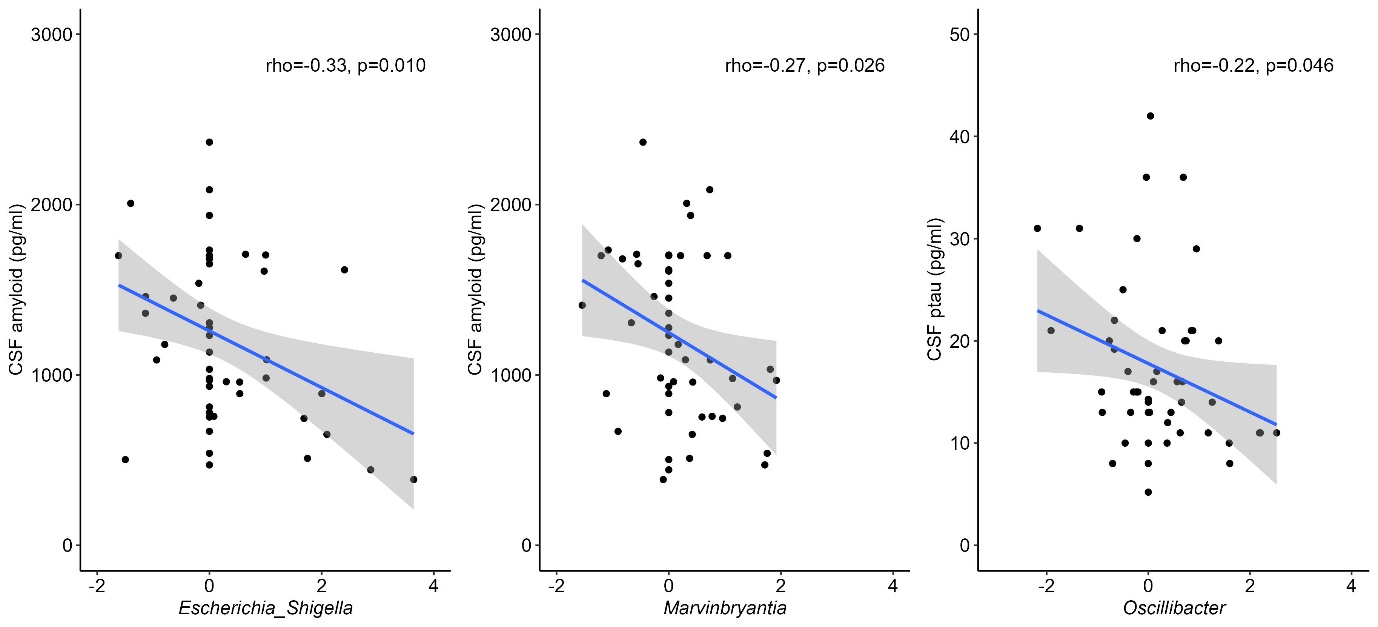

**Figure S6. Spearman’s rank correlation plots bewteen gut microbiota abundance and CSF AD biomarkers.** X axis shows the centered log ratio-transformed counts for each genus. Regression line (in blue) with 95% confidence intervals (gray zone) are shown. Due to missing abundance values for *Escherichia Shigella* (24 of 50 individuals), *Marvinbryantia* (15 of 50 individuals) and *Oscillibacter* (6 of 50 individuals) and prior CLR transformation, we added a pseudocount of 0.0001 in all samples for visualization purposes.

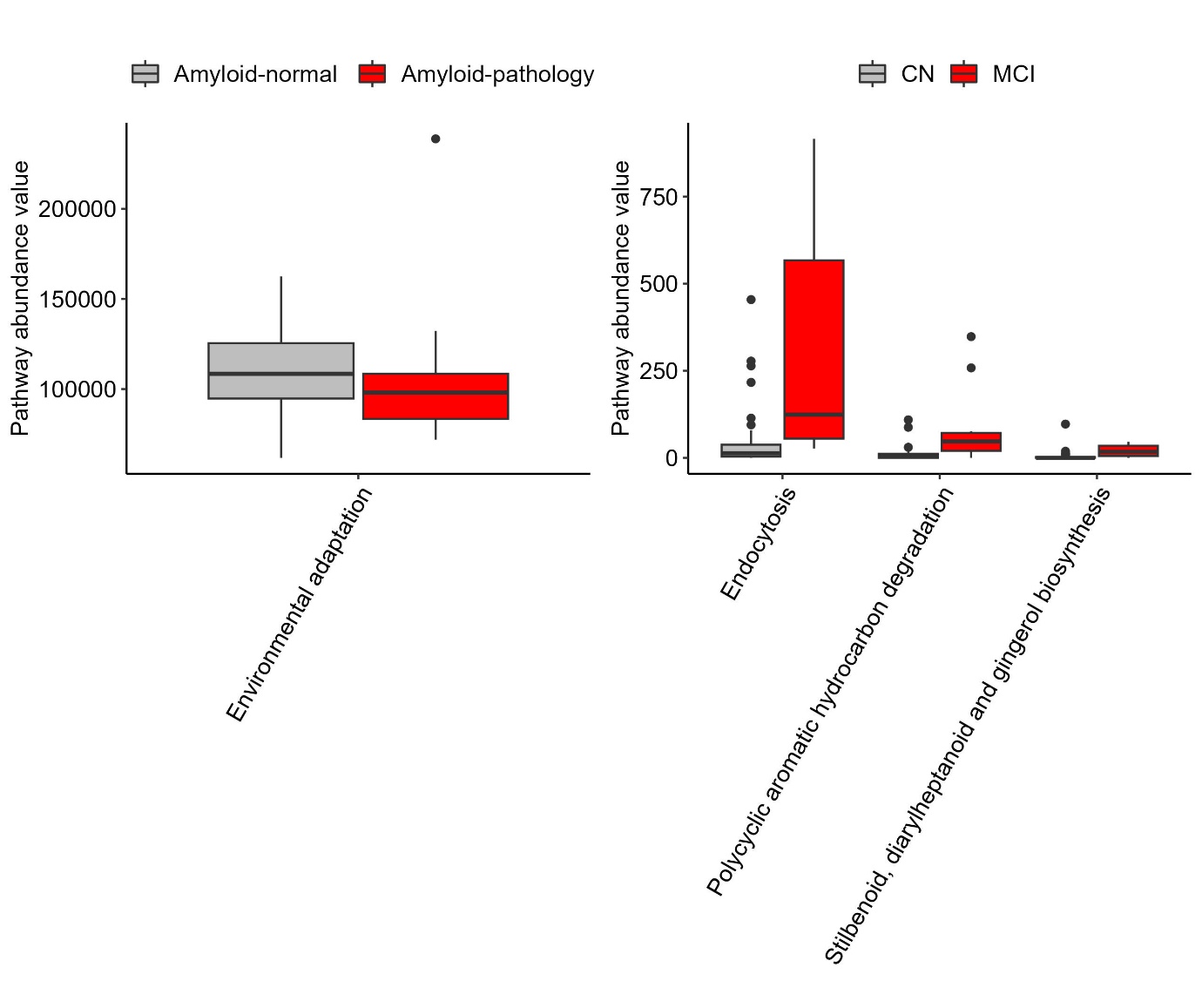

**Figure S7. Functional pathways predicted by Picrust2 that differentiate diagnosis groups.** Left panel shows results for the amyloid-based comparison (2nd KEGG hierarchical level) and right panel shows results for the clinically-based comparison (3rd KEGG hierarchical level). A multivariable logistic regression analysis using LASSO penalty with minimum lambda was applied to detect significant pathways. CN: Cognitively Normal. MCI: Mild Cognitive Impairment.
